# Dose-effect relationship of ginger interposed moxibustion for allergic rhinitis: study protocol for a randomised, placebo-controlled and parallel clinical trial

**DOI:** 10.1101/2021.08.29.21262781

**Authors:** Yunfan Wen, Wen Wang, Yingna Ni, Yeqiao Gui, Zhihai Hu, Yi Wang, Mengke Han, Dongmin Zhang, Shi Shu, Shuang Zhou

**Affiliations:** Shanghai University of Traditional Chinese Medicine, China; Department of Acupuncture, Shanghai TCM-Integrated Hospital, Shanghai University of TCM, China

## Abstract

**Introduction:** Allergic rhinitis has a severe impact on patients’ life quality, and the incidence rate keeps increasing. Moxibustion is widely used for treating allergic rhinitis, and quantity is the basis of moxibustion efficacy. The purpose of this study is to evaluate the relationship between the different quantities of moxibustion and the efficacy of moxibustion in the treatment of allergic rhinitis. This study may be conductive to the standardization of moxibustion and furnish the mechanism of dose-effect relationship of moxibustion with data and new ideas.

**Methods and analysis:** This randomized, placebo-controlled clinical trial will include 33 patients with allergic rhinitis who will be randomly assigned into three groups in a 1:1:1 ratio: high-dose moxibustion group, low-dose moxibustion group and sham moxibustion control group. All groups will be treated once every other day, 20 days for one treatment course. And the patients will receive treatment for 2 courses with an interval of 2 days between courses. We will conduct a follow-up 30 days later after completion of treatments. The primary outcome measure is Total Nasal Symptom Score, carried out at baseline, 3, 6 and 10 weeks. Secondary outcome measure is Rhino Conjunctivitis Quality of Life Questionnaire, carried out at baseline, 6 and 10 weeks.

**Ethics and dissemination:** This trail has been approved by the IRB of Shanghai TCM-Integrated Hospital, Shanghai University of TCM. The results of the trial will be disseminated in peer-reviewed journals.

**Trial registration number:** ChiCTR2100050373; Pre-results.

**Strengths and limitations of this study:**

- A randomised, placebo-controlled and parallel clinical trial will be conducted to test if ginger interposed moxibustion would have curative effect on allergic rhinitis and reveal the preliminary dose-effect relationship of moxibustion.
- This study will set high-dose moxibustion group and low-dose moxibustion group to test if more quantities of ginger interposed moxibustion would achieve better therapeutic effects on allergic rhinitis.
- Further study should be carried out to test if the curative effect of ginger interposed moxibustion would be positively correlated with the quantities.

## INTRODUCTION

Allergic rhinitis (AR) is the most common immunologic disorder worldwide, with more than 500 million people affected. According to statistics, the incidence rate of about 5% to 20%, and the incidence rate keeps increasing.^1^ Allergic rhinitis is a non-infectious inflammation of nasal mucosa induced by IgE after exposure to the allergens.^2^ The major clinical manifestations of allergic rhinitis include sneezing, rhinorrhea, nasal congestion and nasal itching.^3^ Although allergic rhinitis does not threaten patients’ lives, it brings olfaction disorders, nasal symptoms, sleep apnea and so on, affecting patients’ life quality seriously. In addition, allergic rhinitis can cause psychological stress in patients, too.^4^ Allergic rhinitis may lead to otitis media, sinusitis and other diseases.^5^ According to research, medical cost of allergic rhinitis in the United States amounts to $2 billion to $5 billion a year. ^6^

No cure for allergic rhinitis has been found. Treatments to alleviate AR symptoms are mainly pharmacotherapy and immunotherapy. Pharmacotherapy includes oral antihistamines, intranasal glucocorticosteroids and nasal decongestants. Subcutaneous immunotherapy and sublingual immunotherapy are the two types used in immunotherapy. However, both pharmacotherapy and immunotherapy bring undesirable adverse reactions and drug resistance. Pharmacotherapy may cause rhinitissicca, epistaxis, drymouth, dizziness and hypersomnia.^7^ Immunotherapy may lead to rash, pruritus and allergic reactions.^8^

Thus, with the advantages of significant efficacy, rare adverse reactions, low cost, convenience and high acceptance rate, moxibustion has been widely used for treating allergic rhinitis in recent years.^9^ Literatures have reported that moxibustion has advantages in the treatment of allergic rhinitis. ^10-12^ A clinical trial shows that moxibustion can decrease plasma IL-2, IL-6, and cGMP contents and that it can raise plasma cGMP level in allergic rhinitis patients. ^13^ With the photothermal and pharmacological effects, moxibustion can achieve the aims of warming channels to dispel coldness and strengthening healthy qi to eliminate pathogens. ^14^ Since ancient times, the dose-effect relationship of moxibustion has been the focus of doctors of TCM. Quantity is the basis of the effect of moxibustion. Neither overdose nor underdose can achieve the desired effect.^15^ However, there are certain problems existing in clinical trials.^16^ (1) There is a lack of criteria of quantity of moxibustion in the trials of allergic rhinitis. (2) There is a lack of clinical trial on the dose-effect relationship of moxibustion and explanation for the mechanism. (3) There is a lack of observation on the long-term efficacy of moxibustion for allergic rhinitis. The aim of this study is to reveal the dose-effect relationship of moxibustion in treating allergic rhinitis and evaluate its efficacy.

## OBJECTIVES

We will set up ginger interposed moxibustion groups with different quantities of moxibustion and administer the treatment. The purpose is to investigate the efficacy of ginger interposed moxibustion in the treatment of allergic rhinitis. This study will test if more quantities of ginger interposed moxibustion would achieve better therapeutic effects on allergic rhinitis. The trial will reveal the preliminary dose-effect relationship of moxibustion and provide references for the objective development of moxibustion.

## METHODS AND ANALYSIS

### Study design and setting

This randomised, placebo-controlled and parallel clinical trial will be conducted in Shanghai TCM-Integrated Hospital, Shanghai University of TCM. We will enroll patients from the outpatient departments of Acupuncture and Moxibustion in Shanghai TCM-Integrated Hospital, Shanghai University of TCM. The patients will be randomly assigned into three groups in a 1:1:1 ratio:high-dose moxibustion group, low-dose moxibustion group and sham moxibustion control group. Through literature research, we select Dazhui (DU-14), Feishu(BL-13), Pishu (BL-20), Shenshu (BL-23), Shangyintang(extra ordinary point)and Hegu(LI-4)for the study. All groups will be treated once every other day, 20 days for one treatment course. And the patients will receive treatment for 2 courses with an interval of 2 days between courses. We will conduct a follow-up 30 days later after completion of treatments.

We will apply Total Nasal Symptom Score (TNSS) and Rhino Conjunctivitis Quality of Life Questionnaire (RQLQ) to evaluate clinical effects of the three groups. By comparison of TNSS scores and RQLQ of different treatment courses, we will test if ginger interposed moxibustion would be effective in treating AR. And the curative effect will be calculated according to the difference of TNSS scores before and after treatment, which is used to test if the three groups would have statistical difference in effectiveness.

The principle of insulation with wanhua oil: When the moxa cone burns to the bottom soaked with wanhua oil, the moxa cone cannot ignite and will automatically extinguish. Previous studies have completed the preliminary research: the temperature of the bottom of the burning moxa cone soaked with wanhua oil is about 30 °C, the warm feeling temperature of normal human trunk skin is 34-38 °C, and the heat pain feeling temperature is 41-50 °C.^17^ It indicates that moxa cone soaked with wanhua oil will not cause warm stimulation, thus avoiding the special therapeutic effect on human body.

The trial was registered at the China Clinical Trial Registry (ChiCTR1800016371) on 26 August 2021. The study protocol was approved by Shanghai TCM-Integrated Hospital Ethics Committee (2021-051-1). This is the first version of the protocol, 27 August 2021. We will start the study in September, 2021 and is expected to complete by September, 2022. The Standard Protocol Items: Recommendation for Interventional Trials 2013 checklist is shown in online additional file 1. The study flowchart is shown in figure 1.

**Figure 1.**
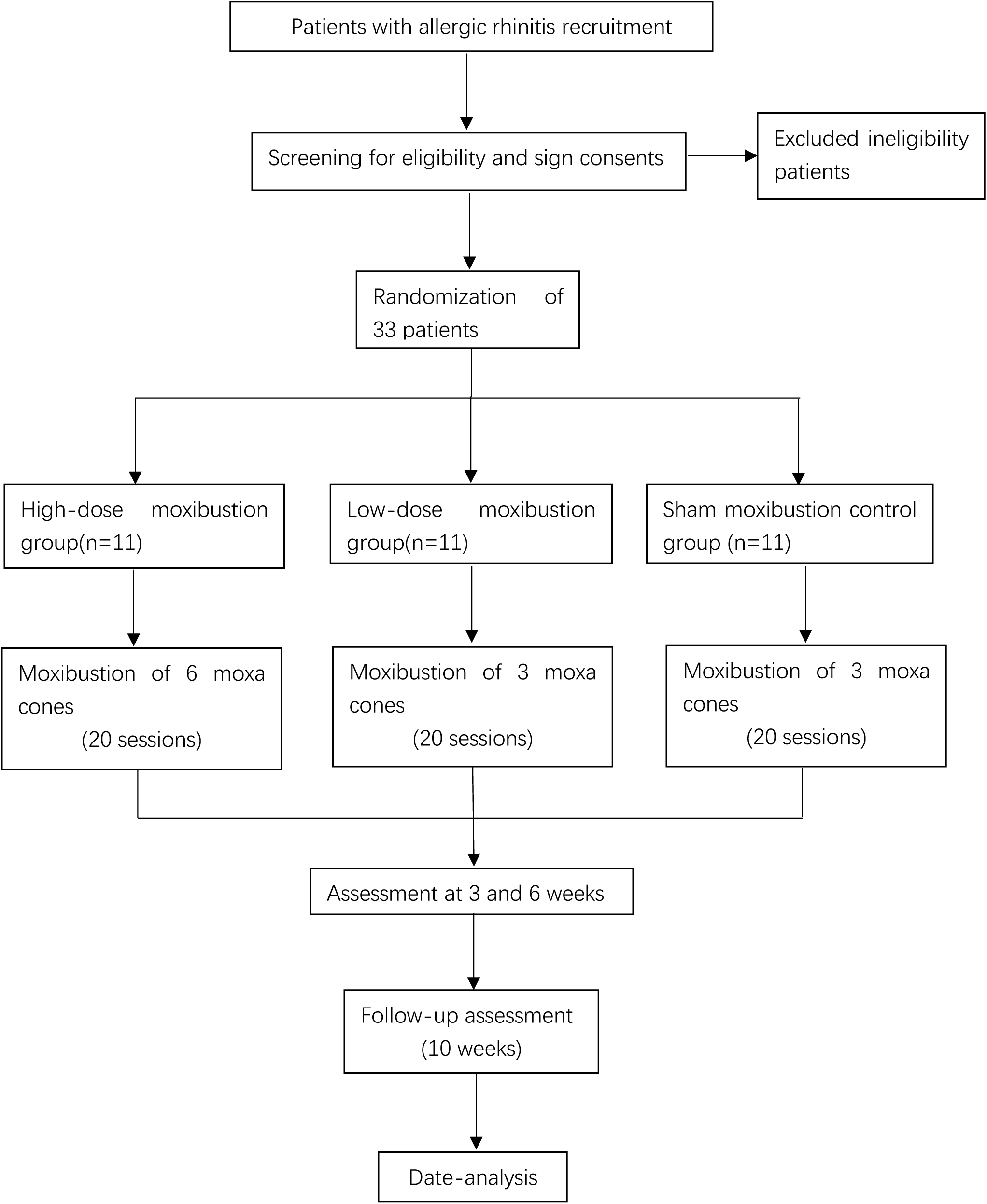
Flowchart of the trial design.

**Figure 2.**
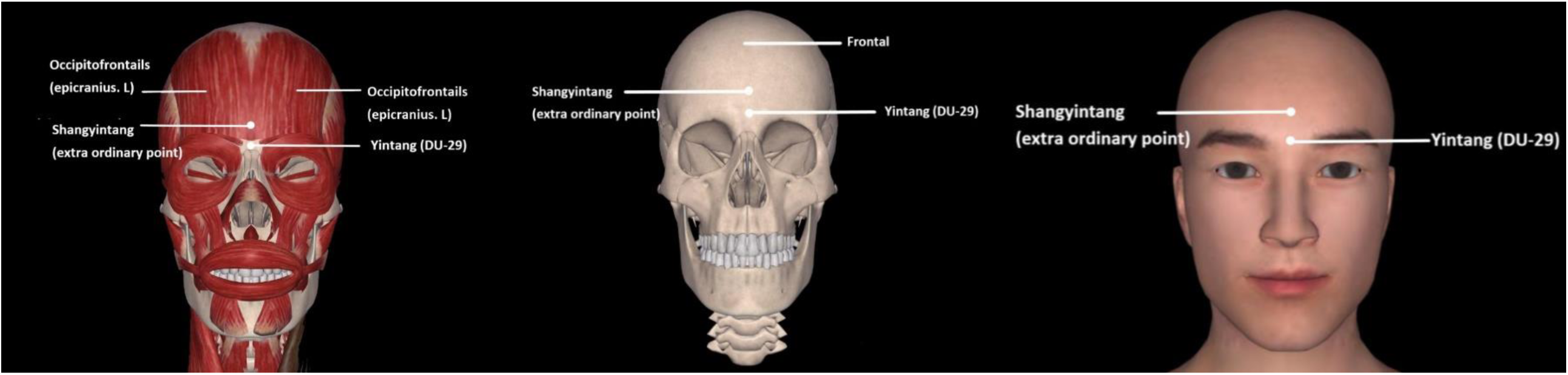
Shangyintang (extra ordinary point)

**Figure 3.**
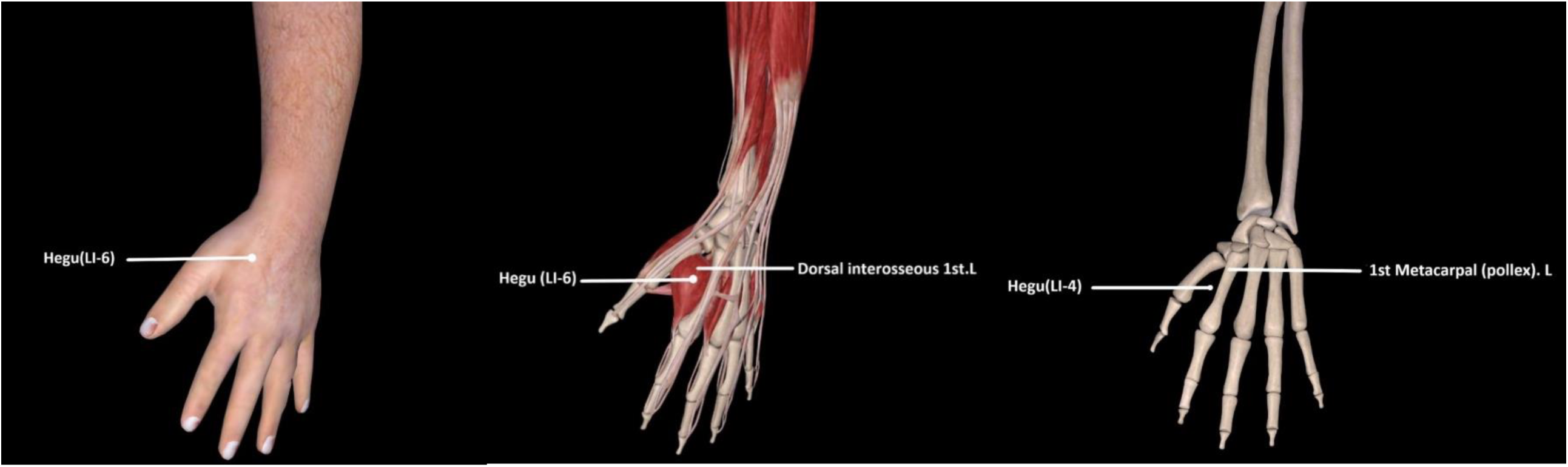
Hegu (LI-4)

**Figure 4.**
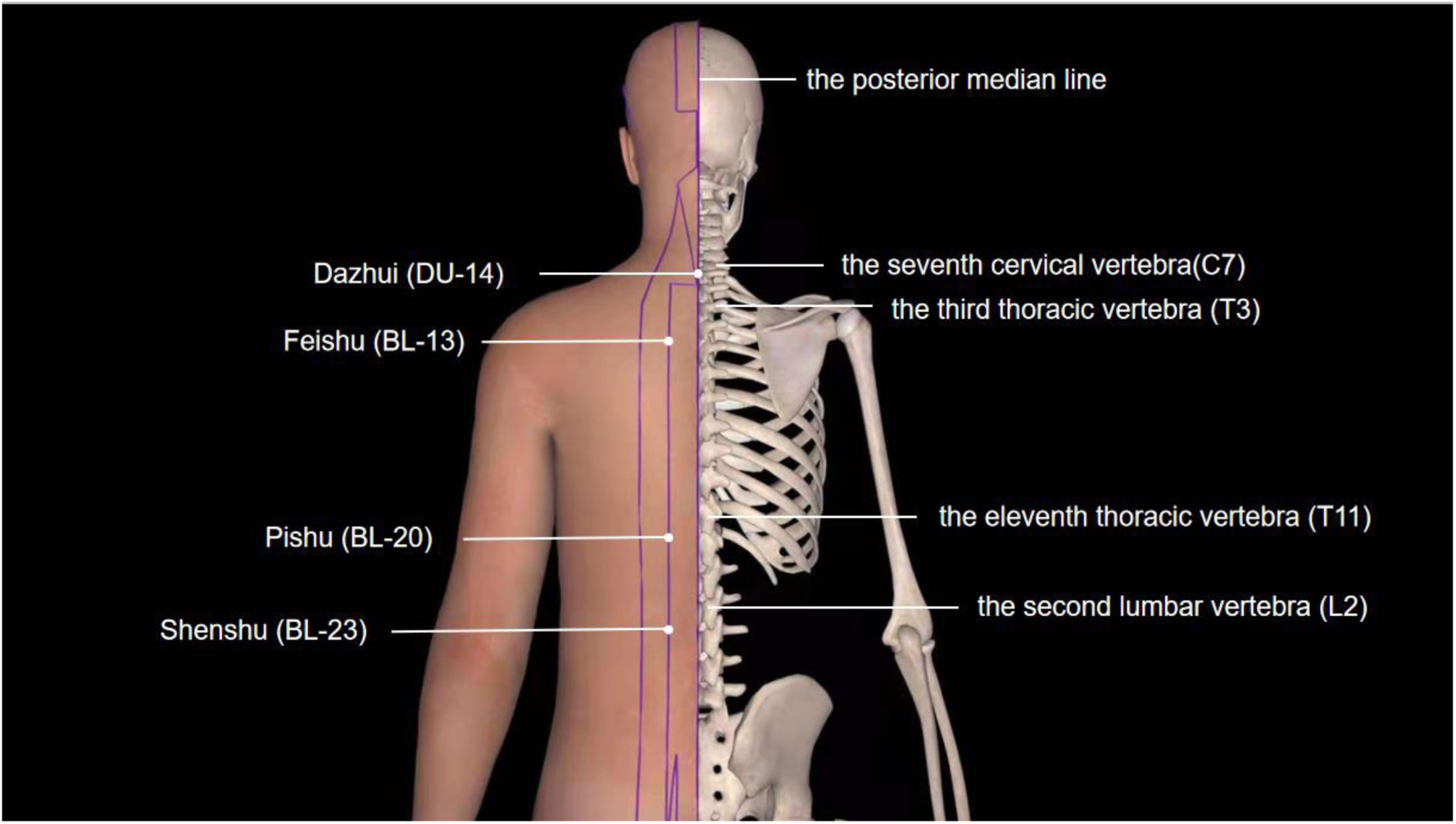
Dazhui (DU-14); Feishu (BL-13); Pishu (BL-20); Shenshu (BL-23).

### Inclusion criteria

The inclusion criteria will be: (1) men or women aged 18–60 years;(2)perennial allergic rhinitis;(3) meet diagnostic criteria of allergic rhinitis;(4) have not used any drugs or treatment methods for allergic rhinitis 2 weeks before the trial;(5) accept ginger moxibustion treatment; (6) obey our advice with treatment;(7) provision of written informed consent for all procedures in this trial.

### Exclusion criteria

Patients will not be enrolled if they meet any of the following:(1) allergic to smoke and dust produced by moxibustion;(2) those who are pregnant, breastfeeding during the study period;(3) serious disease related to the blood vessel, liver, kidney or haematopoietic system;(4) mental illness;(5) complicated with rhinosinusitis, otitis media, nasal polyps, nasal septum deviation and other ENT infections or organic lesions;(6) asthma and other paroxysmal respiratory diseases and in the attack period;(7) not volunteer to participate in this study and refuse to sign informed consent.

### Recruitment and Participants

Recruitment strategies will include posting recruitment advertisements on social media (such as WeChat, which is similar to Facebook) and on notice board in Shanghai TCM-Integrated Hospital, Shanghai University of TCM.

With the assistance of a physician, 33 eligible participants will be identified preliminarily based on the inclusion and exclusion criteria. We will obtain written informed consent from every patient before enrolment. The following criteria are used for diagnosis of allergic rhinitis according to *Chinese guidelines for diagnosis and treatment of allergic rhinitis* formulated by Subspecialty Group of Rhinology (2015), Editorial Board of Chinese Journal of Otorhinolaryngology Head and Neck Surgery: (1) sneezing,(2) runny nose,(3)nasal congestion,(4) nasal itching,(5) eye itching, pharyngeal itching and other symptoms,(6) pale or hyperemia or edema nasal mucosa, and watery secretion in the attack,(7) often induced by contact with allergens or environmental temperature. The diagnosis should meet three of (1, 2,3,4) and (6), and (7).

### Withdrawal criteria and management

Participants who meet one of the following criteria will withdraw from the trial:(1) do not meet the inclusion criteria after inclusion;(2) are poorly compliant or pregnant;(3) voluntarily withdraw from the trail;(4) receive other therapies simultaneously which cause disturbances for measurement of efficacy;(5) occurrence of serious adverse reactions or other unexpected disease changes in the trail.

It is possible for participants to withdraw from the trial due to disease progression, personal intention or serious complications at any time. When participants withdraw, we should know exact reasons and report in CRFs truthfully. If participant does not receive assigned treatment on time, we need ask for exact reasons and follow them up to measure situation after withdrawal.

### Randomization

A statistician will use statistical software SPSS 21.0 to generate random numbers and random distribution tables. Randomization numbers will be placed in sequentially numbered, opaque, sealed envelopes. The random numbers will be arranged in ascending order and be divided into three groups sequentially in a ratio of 1:1:1. The eligible patients will be given envelopes according to their order of enrollment and be assigned to corresponding groups in accordance with generated codes inside envelopes.

### Blinding

After assignment to interventions, the treatment and assessment will be performed independently. Assessor and statistician will be blinded.

### Patients and public involvement

Patients and the public are not involved in designing or conducting the study or the outcome measures.

## INTERVENTIONS

After informed consent is obtained and a baseline evaluation performed, participants will be randomized to three study groups: high-dose moxibustion group, low-dose moxibustion group and sham moxibustion control group. All groups will be treated once every other day, 20 days for one treatment course. And the patients will receive treatment for 2 courses with an interval of 2 days between courses. We will conduct a follow-up 30 days later after completion of treatments. Before the trial researchers will participate in specialized training, by which standard intervention details including manipulation procedures and location of acupoints will be taught to researchers.

During the trial, participants are prohibited from using any drugs or other treatment methods for allergic rhinitis to avoid disturbance for the result of the trial.

### High-dose moxibustion group

Participants will be required to take prone position and expose the back in a fully equipped and temperature controlled room(24-28°C). Fresh slices of ginger (3 cm in diameter and 2-4 mm in thickness with pinpricks) will be placed at 7 standard acupoints: Dazhui(DU-14), bilateral Feishu(BL-13)、Pishu(BL-20) and Shenshu(BL-23)(table 1). Then prepared moxa cones^3^ (2.5 cm in diameter, 2 cm in height and 2.5 g in weight.) will be respectively placed above the ginger and ignited from the bottom. When one moxa cone burns out, replace it with a new moxa cone, with 6 moxa cones per acupoint. After completion of moxibustion in the back part, participants will take supine position. We will perform moxibustion in the same way at acupoints Shangyintang(extra ordinary point), bilateral Hegu(LI-4),with 6 moxa cones per acupoint.

**Table 1.**
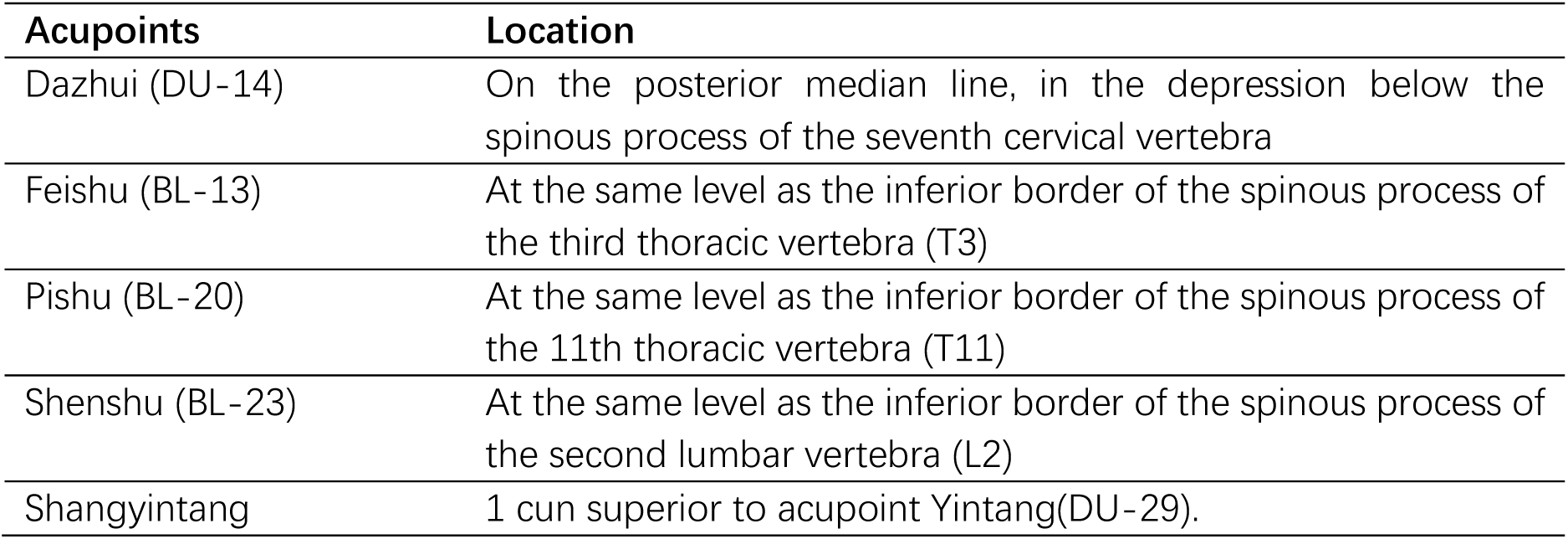
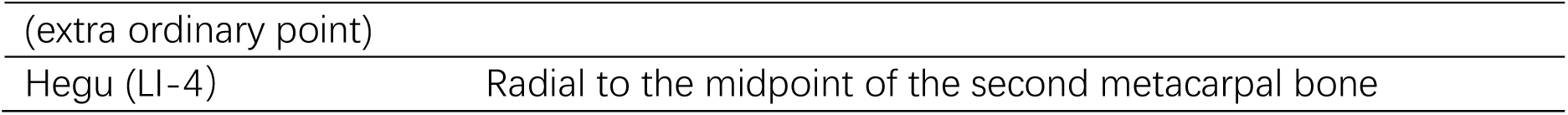
Acupoints selected for use in the study (based on national standard *The Name and Location of Acupoints* (GB/T 12346-2006))

|  |  |
| --- | --- |
| **Acupoints** | **Location** |
| Dazhui (DU-14) | On the posterior median line, in the depression below the spinous process of the seventh cervical vertebra |
| Feishu (BL-13) | At the same level as the inferior border of the spinous process of the third thoracic vertebra (T3) |
| Pishu (BL-20) | At the same level as the inferior border of the spinous process of the 11th thoracic vertebra (T11) |
| Shenshu (BL-23) | At the same level as the inferior border of the spinous process of the second lumbar vertebra (L2) |
| Shangyintang | 1 cun superior to acupoint Yintang(DU-29). |

<sup>3</sup> Moxa cone: Counting unit of moxibustion with amugwort cone

|  |  |
| --- | --- |
| (extra ordinary point) |  |
| Hegu (LI-4) | Radial to the midpoint of the second metacarpal bone |

Every time Shangyintang(extra ordinary point) is performed moxibustion, paper which can fit the forehead will be placed between Shangyintang(extra ordinary point) and participants’ eyes. In this way smoke or smell produced by moxibustion can be blocked to avoid stimulating participant’s nose and eyes. During the treatment, we will maintain a good ventilation and exhaust smoke. If participants feel an unbearable burning sensation, ginger slices will be changed to relieve pain. It is acceptable to move ginger following the line of Du Channel to alleviate burning sensation when performing moxibustion at Shangyintang(extra ordinary point).

### Low-dose moxibustion group

Participants will receive low-dose ginger interposed moxibustion. Intervention procedures will be generally similar to high-dose moxibustion group. The only difference is that each identified acupoint requires only 3 moxa cones.

### Sham moxibustion control group

The intervention procedures for this group will be similar to other two groups. Participants will be given sham moxibustion at the same acupoints. Moxa cones used in this group will be specially processed. Lower 1/3 of moxa cone will be soaked in wanhua oil^4^ for inflaming retarding and heat insulation. ^18^ Then prepared moxa cones will be placed on the ginger slices and ignited from the top, with 3 moxa cones per acupoint.

### Follow-up

Participants will receive treatment for 6 weeks and will be followed up after 4 weeks. At follow-up period, participants will be evaluated through phone calls, WeChat or E-mail. Specific items will be recorded (table 2).

**Table 2.** Measurements to be taken at each point in this trial

|  | Screening | Treatment period |  |  | Follow-up period |
| --- | --- | --- | --- | --- | --- |
| Timepoint | -1 week | 0 week | 3 weeks | 6 weeks | 10 weeks |
| Enrolment |  |  |  |  |  |
| Informed consent form |  | × |  |  |  |
| Inclusion and exclusion criteria | × | × |  |  |  |
| Demographic characteristics | × | × |  |  |  |
| Medical history | × | × | × | × | × |
| Physical examination | × | × | × | × | × |
| Complicating disease | × | × | × | × | × |
| Randomization | × |  |  |  |  |
| Interventions |  |  |  |  |  |
| High-dose moxibustion group |  | 20 sessions of high-dose moxibustion at acupoints |  |  |  |
| Low-dose moxibustion group |  | 20 sessions of low-dose moxibustion at acupoints |  |  |  |
| Sham moxibustion control group |  | 20 sessions of sham moxibustion at acupoints |  |  |  |
| Outcomes |  |  |  |  |  |
| TNSS | × |  | × | × | × |
| RQLQ | × |  |  | × | × |
| Overall therapeutic effect |  |  | × | × | × |
| Safety assessments |  |  |  |  |  |
| Adverse events |  | × | × | × | × |
| Assessment of compliance | × | × | × | × | × |
| TNSS, total nasal symptom score; RQLQ, rhino conjunctivitis quality of life questionnaire. |  |  |  |  |  |

TNSS, total nasal symptom score; RQLQ, rhino conjunctivitis quality of life questionnaire.

### Outcome measurements

#### primary outcomes

The primary outcome is Total Nasal Symptom Score (TNSS)^19^. In accordance with national guidelines, the trial will identify symptoms of AR by using TNSS (total nasal symptom score). TNSS is a valid and reliable instrument that has been used widely in a variety of clinical trials involving patients with allergic rhinitis to evaluate change of nasal symptoms and measure effect of treatment.

We will apply TNSS to evaluate patients’ symptoms: sneezing (the number of continuous sneezes); rhinorrhea (the frequency of blowing nose per day); nasal congestion and nasal itching at baseline and 3, 6 and 10 weeks. The score of every item in TNSS ranges from 0 to 3, where 0=no symptoms, 1=mild symptom(s) (present but bearable), 2=moderate symptom(s) (present and uncomfortable) and 3=severe symptom(s) (unbearable)^20^.

#### secondary outcome

The trial will measure the problems that people with rhino conjunctivitis face in their daily lives by using Rhino Conjunctivitis Quality of Life Questionnaire (RQLQ) ^21^. Previous study has proved that the RQLQ(S) possesses strong discriminative (cross-sectional) and evaluative (longitudinal) measurement properties which make it secure and reliable to measure rhino conjunctivitis quality of life in clinical studies. ^22^

Rhino conjunctivitis Quality of Life Questionnaire (RQLQ) will be applied at baseline and 6 and 10 weeks. It includes 28 questions in seven domains including activity limitations, sleep disturbances, non-nasal or eye symptoms, practical problems, nasal symptoms, ocular symptoms and emotional problems. Participants will be asked to score each question according to their daily experiences and feelings. The score of every item of RQLQ ranges from 0 to 6, with higher scores indicating severer symptoms and more disturbances.

### Overall evaluation of therapeutic effect

Refer to Guidelines for Diagnosis and Treatment of Allergic Rhinitis (2015, Tianjin) formulated by Otorhinolaryngology branch of Chinese Medical Association, the evaluation criteria of therapeutic effect and recurrence condition will be assessed according to TNSS scores at 3, 6 and 10 weeks. Efficacy index(n)= [(preintervention TNSS scores–postintervention TNSS scores)

/preintervention scores]×100%.

Distinctively effective: improvement rate ⩾66%;

Effective: 25<improvement rate <66%;

Ineffective: improvement rate⩽25%.

### Data collection and management

Data will be collected at baseline, 3, 6 weeks and 10 weeks (table 2). The participant’s name, gender, age, course of disease, contact information, medical records and other general information should be recorded prior to trail. To facilitate data reliability, it is indispensable to conduct strict training in standard operating procedure of data collection prior to trail. Researchers will record the case report form (CRF) tables according to participants’ questionnaires to collect data. CRF cannot be casually modified. Any modification needs explanation and should ensure that original record is not covered. The researcher should sign name and date beside the modification. Data in CRF will then be input on an Excel spreadsheet. After verifying accuracy, written CRFs will be locked in cabin and computer files will be stored in a password-protected computer with limited access.

### Safety and moxibustion-related adverse events

It has been reported that moxibustion can cause adverse events such as dizziness, itching, burning sensation, blister, redness, keloids and allergic reactions.23 On each visit, participants are required to report any adverse reactions. Researcher should also observe and document specific symptoms, severity, time, treatment methods, results, relationship with moxibustion in CRFs and report to the chief designer. If serious adverse events are induced, researcher must report to research ethics committee within 24 hours. Participants having serious adverse events should be withdrawn from study and be followed up to receive relevant medical treatment until their condition stabilizes. Connection between adverse events and intervention should be investigated and determined.

### Quality control

Researchers will undergo adequate and professional training before the trial, which help us fully understand protocol and follow guidelines for good clinical practice (GCP), to ensure the safety of patients, adherence to the protocol and correctness of moxibustion intervention. Participant should be filtered in strict accordance with diagnostic criteria, inclusion criteria and exclusion criteria.

### Sample size calculation

According to the clinical results and related literature on the treatment of allergic rhinitis through ginger interposed moxibustion, the sample size was estimated by means of multi-group sample size calculation method. The efficiency of the high-dose group of ginger acupuncture is expected to be 93.3%, the low-dose group is expected to be 89.9%, and the control group is expected to be 27%. Considering a two-sided significance level of 5%, power of 95% and a 20% shedding rate, 33 participants are required with a 1:1:1 group allocation rate. The test level α is set to 0.05,and β to 0.1. The calculation process is as follows:

Calculation formula:n=(Z_1-α’/2_+Z_1-β_)^2×[P_1_(1-P_1_)+ P_2_(1-P_2_)]/(δ _ij_^2),n=max{n _ij_,pairs(I, j)}

α’: value of the division of α;

T: number of comparisons,in this trial T=2; after inquiry of table: Z_1-α’/2_=2.24,Z_1-β_=1.28;

P_1_: the efficiency of the first group;

P_2_: the efficiency of the secondary group;

δ_ij_: the difference of effective rate with clinical significance between any two groups;

n _ij_: the sample size required for each of any two groups.

### Statistical analysis

The outcome analyses will employ the intention to treat (ITT) analysis which will be limited to data collected from participants who receive at least one treatment. Missing data will be compensated through the LOCF. The last observation data will act as the final test results. No interim analysis is planned.

Data analysis will be conducted in a blinded pattern by statisticians by the use of SPSS 21.0 statistical software. Descriptive statistics will be displayed for all the baseline variables and outcome measures. Frequency(percentage)will be used for categorical variables such as gender, and mean±SD for continuous variables including age, disease duration, primary and secondary outcome measures.

Comparison of the baseline characteristics will be analyzed by One-way ANOVA test or Kruskal-Wallis rank sum test for continuous variables and χ2 test or Fisher’s exact test for categorical variables. To compare effectiveness of interventions among three groups, the One-way ANOVA test will be conducted for data with normal distribution and homogeneous variance; The Kruskal-Wallis rank sum test will be for abnormally distributed data and ranked data.

To compare same group of multiple time points, for continuous variables with normal distribution, we will use the paired t-test. When variables are not normal distribution, or variance is not homogeneous or variables are ranked data, Friedman rank sum test will be applied. In all statistical tests, two-sided tests will be used and P<0.05 means statistically significant.

### Rights protection

In this study, we will adopt ginger interposed moxibustion therapy, which has been proved safe and effective in the treatment of allergic rhinitis by previous clinical application and research. This clinical trial conducts informed consent in accordance with the requirements of the *Declaration of Helsinki*. Before each patient enrolls in the trial, it is the responsibility of the researchers to provide him or her with a complete and comprehensive written description of the purpose, procedures, and possible adverse reactions of the trial. Patients should be made aware of their right to withdraw from the trial at any time. Each patient participating in the trial must sign an informed consent form and keep it in the study file.

## ETHICS AND DISSEMINATION

The trial has been registered with the China Clinical Trial Registry (ChiCTR1800016371) and approved by IRB of Shanghai TCM-Integrated Hospital, Shanghai University of TCM (2021-051-1). The trial data will be only used for academic research. This study will help clarify the dose-effect relationship of moxibustion in treating allergic rhinitis. We will publish the results of this study in peer-reviewed journals to ensure widespread dissemination.

## DISCUSSION

The ideal therapeutic effect of moxibustion depends on a certain quantity of moxibustion. And the study of the optimal quantity of moxibustion is conducive to improve the therapeutic effect, reduce the loss of moxibustion materials and improve the acceptance of patients. At present, there is no study on the dose-effect relationship of ginger interposed moxibustion in the treatment of allergic rhinitis. Thus, the purpose of this study is to preliminarily reveal the dose-response relationship of ginger interposed moxibustion in the treatment of AR, and provide scheme reference for clinical care.

Previous studies have shown that ginger interposed moxibustion has good clinical efficacy in the treatment of allergic rhinitis^24-25^, and the operation is simple and easy for patients to adhere to. And “3 moxa cones” and “6 moxa cones” are the doses most used in studies of dose-effect relationship of moxibustion^26 27,28,29^. In this trial, patients will be divided into high-dose moxibustion group (6 moxa cones), low-dose moxibustion group (3 moxa cones) and sham moxibustion group (3 moxa cones). The purpose of setting up sham moxibustion group is to eliminate the placebo effect of moxibustion and verify the effectiveness of moxibustion in the treatment of AR; The two results of high-dose moxibustion group and low-dose moxibustion group will be compared to illustrate the relationship between the quantity and the effect of ginger interposed moxibustion in treating AR.

We expect that moxibustion is effective in the treatment of AR, and the curative effect of high-dose moxibustion group is better than that of low-dose moxibustion group. However, due to the limitation of the objective conditions of the project, this trial will not set more groups of moxibustion volume, so further study should be carried out to test if the curative effect of ginger interposed moxibustion would be positively correlated with the quantities.

## Data Availability

All data will be open to the public one year after finishing the trial.

## Contributors

YW, WW, and YN are responsible for and the cofirst contributors to the concept, design and conduct of the trials and drafting the manuscript. All authors participated in the design of the study and performed the trial. MH and DZ are responsible for recruiting the participants, designing the informed consent and making the picture of acupoints. YG, YW and YN contributed to the writing of the statistical design of the manuscript. WW contributed to the ethical review. All authors read and approved the final manuscript.

## Patient consent for publication

Not required.

## Ethics approval

This trial has been approved by IRB of Shanghai TCM-Integrated Hospital, Shanghai University of TCM (2021-051-1). (June 2021).

## Provenance and peer review

Not commissioned; externally peer reviewed.

## Open access

This is an open access article distributed in accordance with the Creative Commons Attribution Non Commercial (CC BY-NC 4.0) license, which permits others to distribute, remix, adapt, build upon this work non-commercially, and license their derivative works on different terms, provided the original work is properly cited, appropriate credit is given, any changes made indicated, and the use is non-commercial. See: http://creativecommons.org/licenses/by-nc/4.0/.

## Footnotes

3 Moxa cone: Counting unit of moxibustion with amugwort cone

4 Wanhua oil is external analgesic, also called Pain Relieing oil.

## REFERENCES

1 Brozek JL, Bousquet J, Baena-Cagnani CE, et al. Allergic Rhinitis and its Impact on Asthma (ARIA) guidelines: 2010 revision. J Allergy Clin Immunol 2010;126:466–476.

2 Editorial Board of Chinese Journal of Otorhinolaryngology Head and Neck Surgery; Chinese Otorhinolaryngology Society of Chinese Medical Association. Diagnostic and treatment principle for allergic rhinitis and a recommended scheme. Chinese journal of otorhinolaryngology head and neck surgery 2005;40:116–117.

3 Wheatley LM, Togias A. Clinical practice. Allergic rhinitis. N Engl J Med 2015;372:456–63.

4 Pearson DJ. Psychological and somatic interrelationships in allergy and pseudo allergy. Allergy Clin Immunol 1988;81:351.

5 Borres MP. Allergic rhinitis: more than just a stuffy nose. Acta diatrica 2009;98:1088.

6 Seidman MD, Gurgel RK, Lin SY, et al. Clinical practice guideline: allergic rhinitis executive summary. Otolaryngol Head Neck Surg 2015;152:197–206.

7 Zhang DW. Observation on the efficacy of beconase in treating allergic rhinitis. China Prac Med 2009(11):129.

8 Si HF. The observation on clinical effectiveness of warm acupuncture in the treatment of allergic rhinitis. Thesis of master degree, China Academy of Chinese Medical Sciences, 2014.

9 Zhuang JW, Tian YF. Research progress of moxibustion in treating allergic rhinitis. Guangming Journal of Chinese Medicine 2020;35:2266–2270.

10 Wang SY, Zhang J, Du R, et al. Clinical observations on hear-sensitive point moxibustion for perennial allergic rhinitis. Shanghai J Acu-mox 2020;39:734–738.

11 Hu R, Tang S, Liu X, et al. Clinical observation of moxibustion in preventive treatment of seasonal allergic rhinitis. Shanghai J Acu-mox 2020;39:565–569.

12 Liu Z, Ruan Y. Clinical study on yang fire-invigorating moxibustion for allergic rhinitis of lung qi deficiency-cold type. Journal of new Chinese medicine 2019;51:223–225.

13 Ding DM, Li SK, Zhang ZL, et al. Clinical trial of long-snake moxibustion treatment for immune function in allergic rhinitis disease patients. Acupuncture research 2016;41:338–342.

14 Huang KY, Liang S, Sun Z, et al. Startup mechanism of moxibustion warming and dredging function. Chinese Acupuncture & Moxibustion 2017;39:1023–1026.

15 Xie H, Yi SX, Yi Z, et al. The research and thinking on the relationship between moxibustion volume and effectness. Chinese Archives of Traditional Chinese Medicine 2010;28:1003–1005.

16 Yang WH, Zhang SQ, Xue CL. Acupuncture for allergic rhinitis: a systematic review. The Third World Integrative Medicine Congress abstracts. Chinese Association of Integrative Medicine 2007:2.

17 Lu L, Fu JM, Feng FY, et al. Dose-effect relationship in treatment of chronic neck pain with the direct moxibustion of small moxa cone. Chinese Acupuncture & Moxibustion 2019;39(07):734-738+754.

18 Lu L, Fu JM, Feng FY, et al. Dose-effect relationship in treatment of chronic neck pain with the direct moxibustion of small moxa cone. Chinese Acupuncture & Moxibustion 2019;39(07):734-738+754.

19 Cheng L, Dong Z, Kong WJ, et al. Guidelines for Diagnosis and Treatment of Allergic Rhinitis (2015, Tianjin). Chinese Journal of Otolaryngology Head and neck surgery 2016,51 (01): 6–24

20 Kim M-H, Ko Y, Ahn J-H, et al. Efficacy and safety of So-Cheong-Ryong-Tang in treatment of perennial allergic rhinitis: study protocol for a double-blind, randomised, parallel-group, multicentre trial. BMJ Open 2017;7:e016556. doi:10.1136/bmjopen-2017-016556

21 Juniper EF, Guyatt GH. Development and testing of a new measure of health status for clinical trials in rhinoconjunctivitis. Clin Exp Allergy 1991;21:77–83.

22 Juniper EF, Thompson AK, Ferrie PJ, et al. Validation of the standardized version of the Rhinoconjunctivitis Quality of Life Questionnaire. J Allergy Clin Immunol 1999;104:364–9.

23 Park JE, Lee SS, Lee MS, et al. Adverse events of moxibustion: a systematic review. Complement Ther Med 2010;18:215–23.

24 Qin XY, Mi LN, Wu DY. Ginger interposed moxibustion for allergic rhinitis. Chinese Acupuncture & Moxibustion, 2010, 30(10):805.

25 Yang GJ, Liu YL, Xu WG. 60 cases of allergic rhinitis treated by ginger interposed moxibustion at Beishu point. Acupuncture & Moxibustion, 2001(03):20.

26 Lu MX, Wang YS, Wang SC, et al. Experimental observation on the effect of ginger interposed moxibustion with different quantities of moxibustion on local temperature of acupoints in Hyperlipidemia Rats. Journal of Liaoning University of Traditional Chinese Medicine, 2011, 13(02):21–23.

27 Bi DY, Ning JJ, Xu Y, et al. Effects of ginger interposed moxibustion with different quantities on the expression of mitogen extracellular kinase and extracellular regulatory protein kinase in gastric tissue of rats with spleen deficiency syndrome. Chinese Journal of Information on Traditional Chinese Medicine, 2018, 25(01):54–58.

28 Jia LJ, Wang SC, Wang YS, et al. Effects of ginger interposed moxibustion on regulating blood lipid. Chinese Journal of Information on Traditional Chinese Medicine, 2011, 18(01):47–48.

29 Bi DY, Ning JJ, Xu Y, et al. Effects of different doses of ginger-partitioned moxibustion on trefoil factor 1, mucin 5AC and epidermal growth factor receptor in rats with spleen deficiency syndrome. Journal of Acupuncture and Tuina Science, 2018, 16(1):

